# Use of the first National Early Warning Score recorded within 24 hours of admission to estimate the risk of in-hospital mortality in unplanned COVID-19 patients: a retrospective cohort study

**DOI:** 10.1101/2020.07.13.20144907

**Authors:** Donald Richardson, Muhammad Faisal, Massimo Fiori, Kevin Beatson, Mohammed A Mohammed

## Abstract

**Objectives:** Although the National Early Warning Score (NEWS) and its latest version NEWS2 are recommended for monitoring for deterioration in patients admitted to hospital, little is known about their performance in COVID-19 patients. We aimed to compare the performance of the NEWS and NEWS2 in patients with COVID-19 versus those without during the first phase of the pandemic.

**Design:** a retrospective cross-sectional study

**Setting:** Two acute hospitals (Scarborough and York) are combined into a single dataset and analysed collectively.

**Participants:** Adult (>=18 years) non-elective admissions discharged between 11-March-2020 to 13-June-2020 with an index or on-admission NEWS2 electronically recorded within ±24 hours of admission are used to predict mortality at four-time points (in-hospital, 24hours, 48hours, and 72hours) in COVID-19 versus non-COVID-19 admissions.

**Results:** Out of 6480 non-elective admissions, 620 (9.6%) had a diagnosis of COVID-19. They were older (73.3 vs 67.7yrs), more often male (54.7% vs 50.1%), had higher index NEWS (4 vs 2.5) and NEWS2 (4.6 vs 2.8) scores and higher in-hospital mortality (32.1% vs 5.8%). The c-statistics for predicting in-hospital mortality in COVID-19 admissions was significantly lower using NEWS (0.64 vs 0.74) or NEWS2 (0.64 vs 0.74), however these differences reduced at 72hours (NEWS: 0.75 vs 0.81; NEWS2: 0.71 vs 0.81), 48 hours (NEWS: 0.78 vs 0.81; NEWS2: 0.76 vs 0.82) and 24hours (NEWS: 0.84 vs 0.84; NEWS2: 0.86 vs 0.84). Increasing NEWS2 values reflected increased mortality, but for any given value the absolute risk was on average 24% higher (e.g., NEWS2=5: 36% vs 9%).

**Conclusions:** The index or on-admission NEWS and NEWS2 offer lower discrimination for COVID-19 admissions versus non-COVID-19 admissions. The index NEWS2 is not better than the index NEWS. For each value of the index NEWS or index NEWS2, COVID-19 admissions had a substantially higher risk of mortality than non-COVID-19 admissions which reflects the increased baseline mortality risk of COVID-19.

**Article Summary:**

- This study data is from a single NHS Trust and used the index NEWS/NEWS2 scores. The extent to which these findings are generalisable, especially to minority ethnic groups with higher mortality, needs further study.
- Although we found no evidence of NEWS2 as having a superior performance to NEWS, this does suggest that the additional enhancements in NEWS2 are having a limited impact and the underlying reasons need further study.
- NEWS and NEWS2 are repeatedly updated for each patient according to local hospital protocols, and the extent to which changes in NEWS or NEWS2 over time reflect changes in mortality risk needs further study.

## Introduction

The novel coronavirus SARS-CoV-2, which was declared as a pandemic on 11-March 2020, produces the newly identified disease ‘COVID-19’ in patients with symptoms (Coronaviridae Study Group of the International Committee on Taxonomy of Viruses[1]) which has challenged health care systems worldwide.

Patients with COVID-19 admitted to a hospital can develop severe disease with life threatening respiratory and/or multi-organ failure [2,3] with a high risk of mortality in part due to the lack of an effective treatment for the underlying disease in the early phase of the pandemic. Thus, it is recommended that patients at risk of deterioration are referred to critical care. The appropriate early assessment and management of patients with COVID-19 is important in ensuring high-quality care.

In the UK National Health Service (NHS), the patient’s vital signs are monitored and summarised into a National Early Warning Score (NEWS) or its latest iteration (NEWS2)[4]. NEWS is used across the world [4]. NEWS and NEWS2 are calculated from six physiological variables or vital signs—respiration rate, oxygen saturation, temperature, systolic blood pressure, heart rate and level of consciousness (alert, confusion, voice, pain, unresponsive) and also use of supplemental oxygen—which are routinely collected by nursing staff as an integral part of the process of care, usually for all patients, and then repeated thereafter depending on local hospital protocols. NEWS2 includes two oxygen saturation scales (scale 1 and scale 2) and new confusion[5]. NEWS2 points are allocated according to these clinical observations. A higher NEWS2 correlates with a higher chance of deterioration. Gidari et al. [6] evaluated NEWS2 at hospital admission of patients with COVID-19 as a predictor of ICU admission. Furthermore, Kostakis et al [7] investigated association of the last or ultimate recorded NEWS2/NEWS within 24 hours of death or ICU admission in COVID-19 and non-COVID cohorts.

Although NEWS2 is recommended for clinical use in patients with COVID-19 [8], little is known about how NEWS2 performs in practice. In this study, we aimed to compare the performance of NEWS and NEWS2, in unplanned admissions to a teaching hospital during the first phase of the novel coronavirus SARS CoV-2 (COVID-19) pandemic, in predicting in-hospital mortality at four time points (24hours, 48hours,72hours and in-hospital mortality) in COVID-19 versus non-COVID-19 admissions. For all our analyses we use the on-admission or index NEWS2 because this is an early indicator of the severity of illness.

## Methods

### Setting & data

Our cohort of unplanned admissions are from two acute hospitals which are approximately 65 kilometres apart in the Yorkshire & Humberside region of England – Scarborough hospital (n∼300 beds) and York Hospital (YH) (n∼700 beds), managed by York Teaching Hospitals NHS Foundation Trust. For the purposes of this study, the two acute hospitals are combined into a single dataset and analysed collectively. The hospitals have electronic NEWS2 scores and vital signs recording which are routinely collected as part of the patient’s process of care.

We considered all adult (age≥18 years) emergency medical admissions (non-elective/unplanned excluding ambulatory care area patients), discharged during 3 months (11 March 2020 to 13 June 2020), with electronic NEWS2 recorded within ±24 hours of admission. For each emergency admission, we obtained a pseudonymised patient identifier, patient’s age (years), gender (male/female), ethnicity, body mass index (BMI kg/m^2^), discharge status (alive/dead), admission and discharge date and time, diagnoses codes based on the 10th revision of the International Statistical Classification of Diseases (ICD-10) [9] [10], NEWS2 (including its subcomponents respiratory rate, temperature, systolic pressure, pulse rate, oxygen saturation, oxygen supplementation, oxygen scales 1 & 2, and alertness including confusion) [4,5]. The diastolic blood pressure was recorded at the same time as systolic blood pressure. Historically, diastolic blood pressure has always been a routinely collected physiological variable on vital sign charts and is still collected where electronic observations are in place (see Table S1 & S2 in supplementary material). NEWS2 produces integer values that range from 0 (indicating the lowest severity of illness) to 20 (the maximum NEWS2 value possible). The index NEWS2 was defined as the first electronically recorded NEWS2 within ±24 hours of the admission time as vital signs can be collected before admission. We excluded records where the first NEWS2 was not within ±24 hours of admission or was missing/not recorded (see Table 1). Since NEWS2 extends NEWS, we used the same dataset to compare NEWS and NEWS2 especially as NEWS is still in widespread use. The ICD-10 code ‘U071’ was used to identify records with COVID-19. We searched, primary and secondary ICD-10 codes for ‘U071’ for identifying COVID-19.

**Table 1.** Characteristics of emergency medical admissions in COVID-19 versus non-COVID-19.

| Characteristic | COVID-19 | Non-COVID-19 | p-value |
| --- | --- | --- | --- |
| N | 620 | 5824 | - |
| Male (%) | 339 (54.7) | 2918 (50.1) | 0.033 |
| Mean Age [years] (SD) | 73.3 (15.4) | 67.7 (19) | <0.001 |
| **Admission type** |  |  | <0.001 |
| Medical | 588 (94.8) | 4727 (81.1) |  |
| Surgical | 32 (5.2) | 1097 (18.9) |  |
| **Ethnicity** |  |  | <0.001 |
| White | 465 (75) | 4668 (80.2) |  |
| Black, Asian, and other minority ethnic | 34 (5.5) | 152 (2.6) |  |
| Missing | 121 (19.5) | 1004 (17.2) |  |
| Median BMI (IQR)* [kg/m<sup>2</sup>] | 27.5 (8.4) | 26 (7.6) | <0.001 |
| Mean NEWS (SD) | 4 (2.8) | 2.5 (2.3) | <0.001 |
| Mean NEWS2 (SD) | 4.6 (3) | 2.8 (2.7) | <0.001 |
| **Vital Signs** |  |  |  |
| Mean Respiratory rate [breaths per minute] (SD) | 23.5 (6.6) | 19.8 (5) | <0.001 |
| Mean Temperature [oC] (SD) | 36.8 (1.1) | 36.3 (0.9) | <0.001 |
| Mean Systolic pressure [mmHg] (SD) | 136.1 (25.8) | 142.5 (29.2) | <0.001 |
| Mean Diastolic pressure [mmHg] (SD) | 76.5 (16.3) | 79.4 (16.8) | <0.001 |
| Mean Pulse rate [beats per minute] (SD) | 92.2 (22.1) | 88.5 (22.2) | <0.001 |
| Mean % Oxygen saturation (SD) | 94.8 (4.4) | 96.4 (2.9) | <0.001 |
| Oxygen supplementation (%) | 207 (33.4) | 667 (11.5) | <0.001 |
| Mean oxygen flow rate [Litre per minute] (SD) | 7.6 (5.8) | 6.4 (5.5) | 0.008 |
| Oxygen scale 2 (%) | 42 (6.8) | 361 (6.2) | 0.634 |
| **Alertness** |  |  | <0.001 |
| Alert (%) | 514 (82.9) | 5239 (90) |  |
| Baseline confusion (%) | 5 (0.8) | 45 (0.8) |  |
| New confusion (%) | 19 (3.1) | 82 (1.4) |  |
| Pain (%) | 0 (0) | 49 (0.8) |  |
| Voice (%) | 58 (9.4) | 227 (3.9) |  |
| Unconscious (%) | 24 (3.9) | 182 (3.1) |  |
| Referred to critical outreach team (%) | 91 (14.7) | 211 (3.6) | <0.001 |
| Admission to ICU (%) | 42 (6.8) | 147 (2.5) | <0.001 |
| Palliative care (%) | 65 (10.5) | 288 (4.9) | <0.001 |
| On ventilation (%) | 18 (2.9) | 12 (0.2) | <0.001 |
| Median Length of Stay (days) (IQR) | 7.3 (11.7) | 3 (5.5) | <0.001 |
| Mortality with-in 24 hours (%) | 9 (1.5) | 53 (0.9) | 0.273 |
| Mortality with-in 48 hours (%) | 15 (2.4) | 94 (1.6) | 0.189 |
| Mortality with-in 72 hours (%) | 33 (5.3) | 131 (2.3) | <0.001 |
| In-hospital Mortality | 199 (32.1) | 336 (5.8) | <0.001 |

*\* BMI is missing 188 (30.3%) for COVID and 2283 (39.2%) for Non-COVID*

### Statistical Modelling

We began with exploratory analyses including line plots that showed the relationship between age, vital signs, NEWS2/NEWS and risk of in-hospital death in COVID-19 and non-COVID-19. We compared the continuous covariates using a two-sample independent t-test (for normal data) or Wilcoxon rank-sum test (for non-normal data). We compared the categorical covariates using a Chi-square proportion test. P-values less than 0.05 were defined as statistically significant.

We determined the discrimination of NEWS and NEWS2 using the concordance or c-statistic which is interpreted as the probability that a deceased patient has a higher risk of death than a randomly chosen non-deceased patient. For a binary outcome (alive/died), the c-statistic is the area under the Receiver Operating Characteristics (ROC) curve [11]. The ROC curve is a plot of the sensitivity, (true positive rate), versus 1-specificity, (false positive rate), for consecutive predicted risks. A c-statistic of 0.5 is no better than tossing a coin, whilst a perfect model has a c-statistic of 1. In general, values less than 0.7 are considered to show poor discrimination, values of 0.7 to 0.8 can be described as reasonable, and values above 0.8 suggest good discrimination[12]. We developed two separate logistic regression models for predicting in-hospital mortality with NEWS and NEWS2 as covariates respectively. We assessed the performance of the index NEWS or index NEWS2 in predicting the mortality at four specified time points - 24hour, 48hour, 72hour and in-hospital in COVID-19 and non-COVID-19 patients using the c-statistic. For each time point we use the index or on-admission NEWS2/NEWS score.

We assessed the sensitivity, specificity, positive predictive value, negative predictive value and likelihood ratios for NEWS and NEWS2 at values ≥5 which is the usual threshold value for escalation to critical care which equates to a 13% mortality risk under NEWS and an 11% risk under NEWS2. The 95% confidence interval for the c-statistic was derived using DeLong’s method as implemented in the pROC library [13] in R [14]. We followed the STROBE guidelines to report the findings [15]. All analyses were undertaken using R [14] and Stata [16].

### Ethical Approval

This study used de-identified data and received ethical approval from the Health Research Authority (HRA) and Health and Care Research Wales (HCRW) (reference number 19/HRA/0548).

### Patient and Public Involvement

There was no patient involvement in this study.

## Results

### Cohort description

There were 6480 discharges over 3 months. We excluded 36 (0.6%) records because the index NEWS2 was not recorded within ±24 hours of the admission date/time or NEWS2 was missing or not recorded at all (see Table S3 in supplementary material).

We analysed data from 6444 admissions, of which 9.6% (620/6444) were diagnosed COVID-19. The demographic, vital signs and outcome profiles of the COVID-19 versus non-COVID-19 admissions is shown in Table 1 and Figure S1. COVID-19 admissions were older (73.3 vs 67.7, p<0.001), more likely to be male (54.7% vs 50.1%, p<0.001), with higher BMI (kg/m2) (27.5 vs 26, p<0.001) than non-COVID-19 admissions. Furthermore, they had higher index NEWS (4.0 vs 2.5, p<0.001) and index NEWS2 (4.6 vs 2.8, p<0.001) than non-COVID-19 admissions which was reflected in differences in vital signs notably, a higher respiratory rate (23.5 vs 19.8, p<0.001), lower oxygen saturation (94.8% vs 96.4%, p<0.001), higher frequency of oxygen supplementation (33.4% vs 11.5%, p<0.001), lower systolic blood pressure (136.1 mmHg vs 142.5 mm Hg, p<0.001) and less likely to be alert (82.9% vs 90%, p<0.001).

COVID-19 admissions were more likely to be referred to the critical outreach team (14.7% vs 3.6%, p<0.001), admitted to the intensive care unit (ICU) (6.8% vs 2.5%) and referred to palliative care (10.5% vs 4.9%). They also had longer hospital stay (7.3 days vs 3.0 days, p<0.001) and higher in-hospital mortality (32.1% vs 5.8%, p<0.001).

Figure 1 shows the relationship between continuous covariates and the observed risk of in-hospital mortality in COVID-19 versus non-COVID-19 admissions. Whilst the pattern of mortality was broadly similar between COVID-19 and non-COVID-19 admissions, COVID-19 admissions had a consistently higher risk of mortality for the range of covariate values (see Figure 1 and Figure S2 in supplementary material). Figure 1 also shows that although increasing NEWS and NEWS2 scores reflected increased mortality, but for any given value of NEWS or NEWS2 the risk of mortality for COVID-19 was on average 24% higher and at a NEWS or NEWS2 of 5 the risk of mortality in COVID-19 vs Non-COVID-19 was 36% versus 9%.

**Figure 1.**
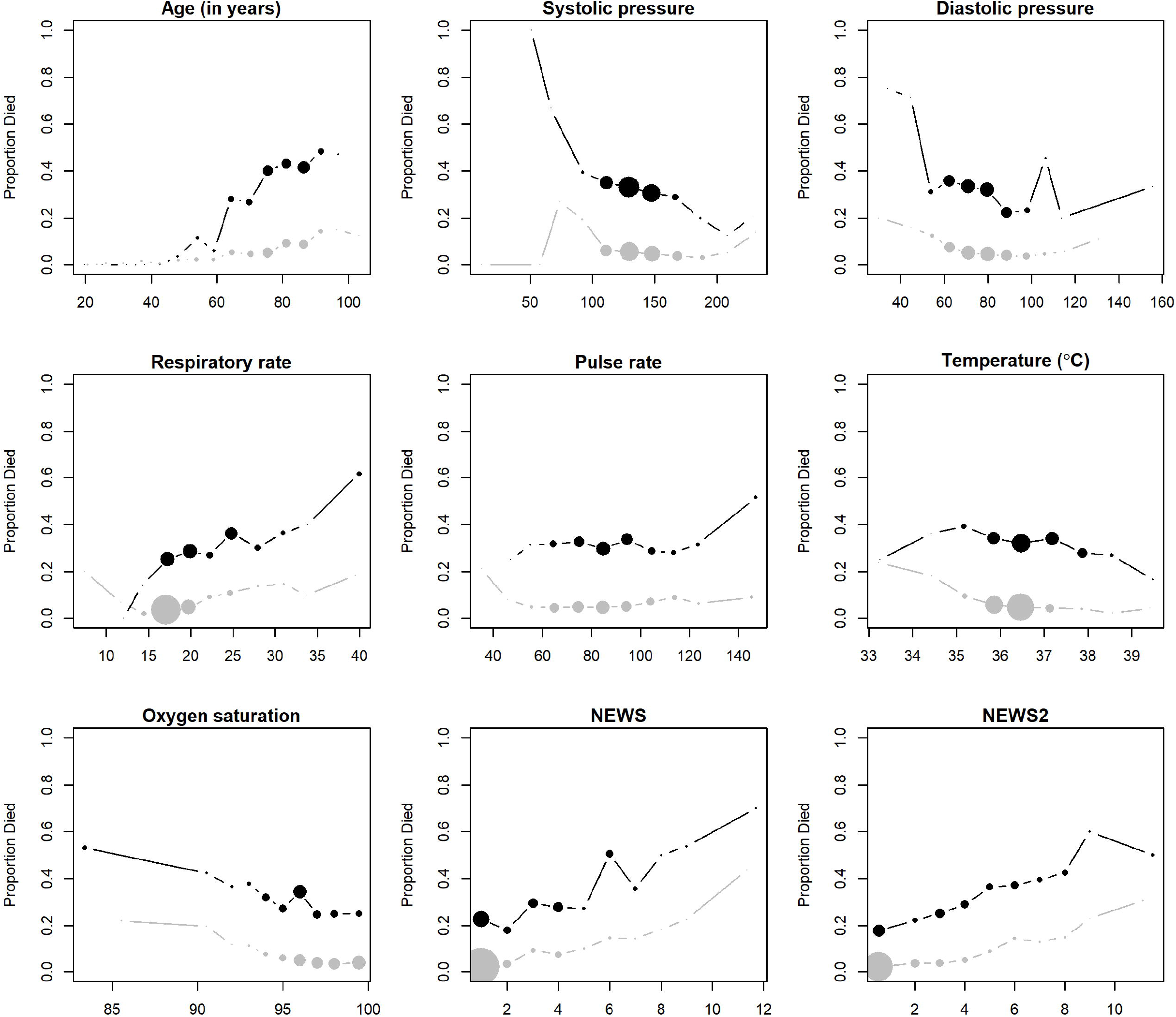
Line plots showing the observed risk of in-hospital mortality with continuous covariates for COVID-19 (black colour) and Non-COVID-19 (grey colour) admissions. Note: Size of circles reflects sample size independently in the COVID-19 and non-COVID-19 groups.

The performance of index NEWS2 to predict the risk of death (24hour, 48hour, 72hour, in-hospital) in COVID-19 and non-COVID-19 admissions is shown in Figure 2 and Table S4. The c-statistics for predicting in-hospital mortality in COVID-19 admissions was significant lower than for patients without COVID-19 (NEWS: 0.64 vs 0.74; NEWS2: 0.64 vs 0.74), however these differences reduced at 72hours (NEWS: 0.75 vs 0.81; NEWS2: 0.71 vs 0.81), 48 hours (NEWS: 0.78 vs 0.81; NEWS2: 0.76 vs 0.82) and 24hours (NEWS: 0.84 vs 0.84; NEWS2: 0.86 vs 0.84). We found the same performance for medical and surgical admission (see Table S5 in supplementary material).

**Figure 2.**
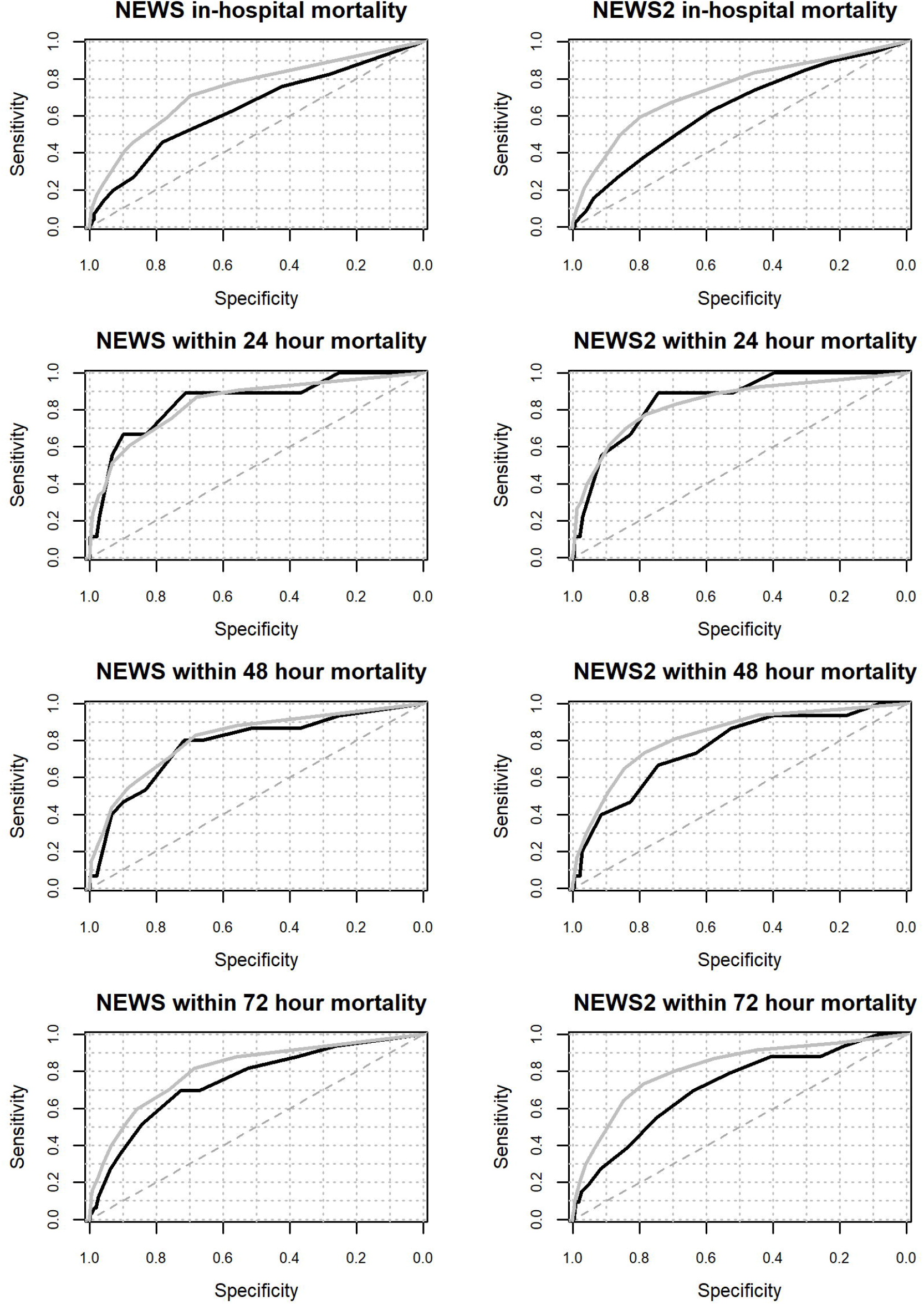
Receiver Operating Characteristic curve for NEWS2 and NEWS in predicting the risk of in-hospital mortality, mortality within 24 hours, 48 hours, and 72 hours in the COVID-19 (black colour) and Non-COVID-19 (grey colour) admissions.

Table 2 includes the sensitivity, specificity, positive and negative predictive values for NEWS and NEWS2 for COVID-19 and non-COVID-19 patients. NEWS2 had higher sensitivity but lower specificity compared to NEWS.

**Table 2.** Sensitivity analysis of NEWS versus NEWS2 in predicting the risk in-hospital mortality, mortality within 24 hours, 48 hours, and 72 hours at NEWS (or NEWS2)≥5 in the COVID-19 and Non-COVID-19 medical admissions. PPV=Positive Predictive Value; NPV= Negative Predictive Value; LR+=Positive Likelihood Ratio; LR-=Negative Likelihood Ratio; N*= Number of positive cases identified by model at NEWS (or NEWS2)≥5.

| Mortality type | Models | COVID-19 |  |  |  |  |  |  | Non-COVID-19 |  |  |  |  |  |  |
| --- | --- | --- | --- | --- | --- | --- | --- | --- | --- | --- | --- | --- | --- | --- | --- |
|  |  | N* | Sensitivity% | Specificity% | PPV | NPV | LR+ | LR- | N* | Sensitivity% | Specificity% | PPV | NPV | LR+ | LR- |
| In-hospital | NEWS | 183 | 45.7<br>(38.7 to 52.9) | 78.1<br>(73.9 to 82) | 49.7<br>(42.3 to 57.2) | 75.3<br>(71 to 79.3) | 2.1<br>(1.7 to 2.6) | 0.7<br>(0.6 to 0.8) | 710 | 40.8<br>(35.5 to 46.2) | 89.6<br>(88.7 to 90.4) | 19.3<br>(16.5 to 22.4) | 96.1<br>(95.5 to 96.6) | 3.9<br>(3.4 to 4.5) | 0.7<br>(0.6 to 0.7) |
| In-hospital | NEWS2 | 300 | 62.8<br>(55.7 to 69.5) | 58.4<br>(53.6 to 63.2) | 41.7<br>(36 to 47.5) | 76.9<br>(71.9 to 81.4) | 1.5<br>(1.3 to 1.8) | 0.6<br>(0.5 to 0.8) | 1300 | 59.2<br>(53.8 to 64.5) | 79.9<br>(78.9 to 81) | 15.3<br>(13.4 to 17.4) | 97<br>(96.4 to 97.5) | 3<br>(2.7 to 3.3) | 0.5<br>(0.4 to 0.6) |
| Within 24 hours | NEWS | 183 | 88.9<br>(51.8 to 99.7) | 71.4<br>(67.6 to 74.9) | 4.4<br>(1.9 to 8.4) | 99.8<br>(98.7 to 100) | 3.1<br>(2.4 to 4) | 0.2<br>(0 to 1) | 710 | 60.4<br>(46 to 73.5) | 88.3<br>(87.4 to 89.1) | 4.5<br>(3.1 to 6.3) | 99.6<br>(99.4 to 99.7) | 5.1<br>(4.1 to 6.5) | 0.4<br>(0.3 to 0.6) |
| Within 24 hours | NEWS2 | 300 | 88.9<br>(51.8 to 99.7) | 52.2<br>(48.2 to 56.2) | 2.7<br>(1.2 to 5.2) | 99.7<br>(98.3 to 100) | 1.9<br>(1.5 to 2.4) | 0.2<br>(0 to 1.4) | 1300 | 77.4<br>(63.8 to 87.7) | 78.2<br>(77.1 to 79.2) | 3.2<br>(2.3 to 4.3) | 99.7<br>(99.5 to 99.9) | 3.5<br>(3 to 4.1) | 0.3<br>(0.2 to 0.5) |
| Within 48 hours | NEWS | 183 | 80<br>(51.9 to 95.7) | 71.7<br>(68 to 75.3) | 6.6<br>(3.4 to 11.2) | 99.3<br>(98 to 99.9) | 2.8<br>(2.1 to 3.8) | 0.3<br>(0.1 to 0.8) | 710 | 54.3<br>(43.7 to 64.6) | 88.5<br>(87.6 to 89.3) | 7.2<br>(5.4 to 9.3) | 99.2<br>(98.9 to 99.4) | 4.7<br>(3.9 to 5.8) | 0.5<br>(0.4 to 0.6) |
| Within 48 hours | NEWS2 | 300 | 86.7<br>(59.5 to 98.3) | 52.6<br>(48.5 to 56.6) | 4.3<br>(2.3 to 7.3) | 99.4<br>(97.8 to 99.9) | 1.8<br>(1.5 to 2.3) | 0.3<br>(0.1 to 0.9) | 1300 | 73.4<br>(63.3 to 82) | 78.5<br>(77.4 to 79.6) | 5.3<br>(4.2 to 6.7) | 99.4<br>(99.2 to 99.6) | 3.4<br>(3 to 3.9) | 0.3<br>(0.2 to 0.5) |
| Within 72 hours | NEWS | 183 | 69.7<br>(51.3 to 84.4) | 72.7<br>(68.9 to 76.3) | 12.6<br>(8.1 to 18.3) | 97.7<br>(95.8 to 98.9) | 2.6<br>(2 to 3.3) | 0.4<br>(0.2 to 0.7) | 710 | 52.7<br>(43.8 to 61.5) | 88.7<br>(87.9 to 89.6) | 9.7<br>(7.6 to 12.1) | 98.8<br>(98.4 to 99.1) | 4.7<br>(3.9 to 5.6) | 0.5<br>(0.4 to 0.6) |
| Within 72 hours | NEWS2 | 300 | 78.8<br>(61.1 to 91) | 53.3<br>(49.2 to 57.4) | 8.7<br>(5.7 to 12.4) | 97.8<br>(95.5 to 99.1) | 1.7<br>(1.4 to 2.1) | 0.4<br>(0.2 to 0.8) | 1300 | 73.3<br>(64.8 to 80.6) | 78.9<br>(77.8 to 79.9) | 7.4<br>(6 to 8.9) | 99.2<br>(98.9 to 99.5) | 3.5<br>(3.1 to 3.9) | 0.3<br>(0.3 to 0.5) |

## Discussion

Whilst NEWS and NEWS2 are recommended for monitoring patients with COVID-19, we found that the index or on-admission NEWS2/NEWS offer lower discrimination for COVID-19 patients versus non-COVID-19 patients. We also found that the index NEWS2 was not better than index NEWS. For each value of the index NEWS2/NEWS, COVID-19 patients had a substantially higher risk of in-hospital mortality than non-COVID-19 patients, which equated to an average 24% risk difference which reflects the higher baseline risk of mortality in our COVID-19 patients. However, the c-statistics for the index NEWS2/NEWS2 improved with shorter time horizons with the highest discrimination (above 0.8) being seen for predicting mortality risk within 24hours of the index NEWS2/NEWS.

A recent paper by Kostakis et al [7], found good discrimination for NEWS or NEWS2 (c-statistics 0.842-0.894) concluding that their “results support the national and international recommendations for the use of NEWS or NEWS2 for the assessment of acute-illness severity in patients with COVID-19.” In contrast to our approach, Kostakis et al [7] used the last or ultimate recorded NEWS2/NEWS within 24 hours of death or ICU admission. We note that when we consider death within 24 hours of admission, our reported c-statistics for index NEWS2/NEWS are comparable with those of Kostakis et al [7].

So taken together these findings indicate that care must be taken not to interpret the predictive power of the ultimate NEWS or NEWS2 score (taken within 24 hours of death) as being equivalent to the predictive power of the index NEWS of NEWS2 score (or preceding NEWS or NEWS2 scores) for risk of in-hospital mortality. The ultimate NEWS or NEWS2 is an accurate predictor of mortality (plus ICU admission in the case of Kostakis et al) for COVID-19 patients but offers a maximum of 24hours for appropriate interventions. This good performance is less surprising when we note that, with the exception of patients who are characterised by abnormal physiology (patients recovering from end-stage renal failure or patients recovering from brain injury), “Patients die not from their disease but from the disordered physiology caused by the disease.” [17]. But, as our findings show, the performance of the index NEWS or index NEWS2 for predicting death in hospital, which offers an early window of opportunity for assessment and intervention, is poorer especially for COVID-19 patients. This needs to be brought to the attention of medical and nursing staff and reflected in escalation protocols and guidelines (which have always highlighted the importance of clinical judgement) to mitigate potential threats to patient safety by promoting situational awareness about the actual, on admission, in-hospital mortality risk for COVID-19 patients.

The World Health Organisation (WHO) describes the range of symptoms seen in COVID-19 which include (but are not limited to) dyspnoea, reduced alertness, delirium, fever, tachypnoea and hypoxia (as a common sign in moderate to severe disease). These symptoms are included in the physiological observation set underpinning NEWS and NEWS2 and were more frequent in our COVID-19 patients compared to non-COVID-19 patients. We also found evidence of lower blood pressure and a higher pulse rate in COVID-19 patients. The NEWS2 guidelines[8] do note that patients with COVID-19 can develop ‘silent hypoxia’ where oxygen saturations can drop to low levels and precipitate acute respiratory failure quickly without the presence of obvious symptoms of respiratory distress. As such any patients admitted and on supplemental oxygen may develop a rapidly increasing oxygen requirement that may not increase the NEWS2 score. It is stressed that any increase in oxygen requirement should trigger an escalation for review by a competent senior decision-maker [8].

Consideration should be also be given to enhancing NEWS or NEWS2 so that they can be used in COVID-19 and non-COVID-19 patients rather than needing to change scoring systems or adjust estimations of risk dependent on diagnosis. We have previously demonstrated how a fully automated computer-enhanced NEWS score can be developed which requires no additional data collection and builds on the standardisation provided by NEWS [18]. We now propose to extend this to include COVID-19 status.

There are several limitations to our study. (1) This study data is from a single NHS Trust and the extent to which these findings are generalisable, especially to minority ethnic groups with higher COVID-19 mortality, needs further study. (2) We used the index NEWS2 which reflects the ‘on-admission’ risk of mortality of the patients. Nonetheless, NEWS2 is repeatedly updated for each patient according to local hospital protocols, and the extent to which changes in NEWS2 over time reflect changes in mortality risk needs further study. (3) Although we found no evidence of NEWS2 as having a superior performance to NEWS, it is important to note that our index NEWS data are hypothetical in the sense that the Trust has been using NEWS2 since April 2019. Nevertheless, it is worth noting that a recent, albeit small Italian study based on 71 hospitalised COVID-19 patients found NEWS2 to be a good predictor (with a high c-statistic 0.90) of subsequent ICU admission for COVID-19 patients but was not able to consider mortality because of insufficient events [6]. Our study did not consider ICU admissions as an outcome because the number of ICU admissions were low but Kostakis et al [7] used it as a composite outcome with in-hospital mortality (5).

## Conclusions

The index or on-admission NEWS and NEWS2 offer lower discrimination for COVID-19 admissions versus non-COVID-19 admissions. The index NEWS2 is not better than the index NEWS. For each value of the index NEWS or index NEWS2, COVID-19 admissions had a substantially higher risk of mortality than non-COVID-19 admissions which reflects the increased baseline mortality risk of COVID-19.

## Data Availability

Our data sharing agreement with NHS York hospital trust does not permit us to share this data with other parties. Nonetheless if anyone is interested in the data, then they should contact the R&D offices in the first instance.

## Contributorship

DR & MAM had the original idea for this work. MF undertook the statistical analyses with guidance from MAM. MFi & KB extracted the necessary data frames. DR gave a clinical perspective. DR, MF and MAM wrote the first draft of this paper and all authors subsequently assisted in redrafting and have approved the final version.

