## supplementary material for "Use of the first National Early Warning Score recorded within 24 hours of admission to estimate the risk of in-hospital mortality in unplanned COVID-19 patients: a retrospective cohort study"

Table S1: NEWS scoring chart

| **Physiological Parameters** | **3** | **2** | **1** | **0** | **1** | **2** | **3** |
| --- | --- | --- | --- | --- | --- | --- | --- |
| **Respiration Rate** | ≤8 |  | 9 - 11 | **12 - 20** |  | 21 - 24 | ≥25 |
| **Oxygen Saturations** | ≤91 | 92 - 93 | 94 - 95 | **≥96** |  |  |  |
| **Any Supplemental Oxygen** |  | Yes |  | **No** |  |  |  |
| **Temperature** | ≤35.0 |  | 35.1 - 36.0 | **36.1 - 38.0** | 38.1 - 39.0 | ≥39.1 |  |
| **Systolic BP** | ≤90 | 91 - 100 | 101 - 110 | **111 - 219** |  |  | ≥220 |
| **Heart Rate** | ≤40 |  | 41 - 50 | **51-90** | 91 - 110 | 111 - 130 | ≥131 |
| **Level of Consciousness** |  |  |  | **Alert** |  |  | Voice, Pain, or Unconscious |

Table S2: NEWS2 scoring chart

| **Physiological Parameters** | **3** | **2** | **1** | **0** | **1** | **2** | **3** |
| --- | --- | --- | --- | --- | --- | --- | --- |
| **Respiration Rate** | ≤8 |  | 9 - 11 | **12 - 20** |  | 21 - 24 | ≥25 |
| **SpO2 Scale 1 (%)** | ≤91 | 92 - 93 | 94 - 95 | **≥96** |  |  |  |
| **SpO2 Scale 2 (%)** | ≤83 | 84 - 85 | 86 - 87 | **88 - 92**  **≥93 on Air** | 93 – 94 on oxygen | 95 – 96 on oxygen | ≥97 on oxygen |
| **Oxygen Saturations** | ≤91 | 92 - 93 | 94 - 95 | **≥96** |  |  |  |
| **Air or oxygen?** |  | Oxygen |  | **Air** |  |  |  |
| **Temperature** | ≤35.0 |  | 35.1 - 36.0 | **36.1 - 38.0** | 38.1 - 39.0 | ≥39.1 |  |
| **Systolic BP** | ≤90 | 91 - 100 | 101 - 110 | **111 - 219** |  |  | ≥220 |
| **Heart Rate** | ≤40 |  | 41 - 50 | **51-90** | 91 - 110 | 111 - 130 | ≥131 |
| **Level of Consciousness** |  |  |  | **Alert** |  |  | Voice, Pain, Confusion, or Unconscious |

The NEWS [https://www.rcplondon.ac.uk/projects/outputs/national-early-warning-score-news] is based on a scoring system in which a score is allocated to vital signs physiological measurements already undertaken when patients present to or are being monitored in hospital. A score is allocated to each as they are measured, the magnitude of the score reflecting how extreme the parameter varies from the norm. This score is then aggregated, and uplifted for people requiring oxygen.

**Table S3 Number of emergency medical admissions included/excluded**

| **Characteristic** | **COVID-19** | **Non-COVID-19** | **All** |
| --- | --- | --- | --- |
|  | N (%) | N (%) | N (%) |
| Total emergency medical discharges between  11 Mar 20 to 13 June 20 | 622 (9.6%) | 5858 (90.4%) | 6480 (100%) |
| Excluded: No NEWS recorded (%) | 0 (0.0) | 19 (0.3) | 19 (0.3) |
| Excluded: First NEWS after 24 hours of admission (%) | 2 (0.3) | 15 (0.3) | 17 (0.3) |
| Total excluded (%) | 2 (0.3) | 34 (0.6) | 36 (0.6) |
| Total included (%) | 620 (99.7) | 5824 (99.4) | 6444 (99.4) |

| **Mortality timepoint** |  | **COVID-19** | | | | **Nov-COVID-19** | | | |
| --- | --- | --- | --- | --- | --- | --- | --- | --- | --- |
|  | **Model** | **Risk discharged alive** | **Risk discharged**  **deceased** | **Risk Difference** | **AUC**  **(95% CI)** | **Risk discharged alive** | **Risk discharged deceased** | **Risk Difference** | **AUC**  **(95% CI)** |
| In-hospital Mortality | NEWS | 0.11 | 0.16 | 0.06 | 0.64  (0.59 to 0.69) | 0.07 | 0.17 | 0.09 | 0.74  (0.71 to 0.77) |
|  | NEWS2 | 0.11 | 0.15 | 0.04 | 0.64  (0.59 to 0.68) | 0.07 | 0.16 | 0.09 | 0.74  (0.71 to 0.77) |
| Mortality with-in 24 hours | NEWS | 0.12 | 0.32 | 0.20 | 0.84  (0.7 to 0.99) | 0.08 | 0.25 | 0.17 | 0.84  (0.78 to 0.89) |
|  | NEWS2 | 0.12 | 0.28 | 0.16 | 0.86  (0.75 to 0.97) | 0.08 | 0.24 | 0.16 | 0.84  (0.78 to 0.9) |
| Mortality with-in 48 hours | NEWS | 0.12 | 0.26 | 0.14 | 0.78  (0.65 to 0.91) | 0.08 | 0.22 | 0.14 | 0.81  (0.77 to 0.86) |
|  | NEWS2 | 0.12 | 0.23 | 0.11 | 0.76  (0.64 to 0.89) | 0.08 | 0.21 | 0.14 | 0.82  (0.78 to 0.87) |
| Mortality with-in 72 hours | NEWS | 0.12 | 0.23 | 0.11 | 0.75  (0.66 to 0.84) | 0.08 | 0.21 | 0.13 | 0.81  (0.77 to 0.85) |
|  | NEWS2 | 0.12 | 0.21 | 0.09 | 0.71  (0.62 to 0.8) | 0.08 | 0.21 | 0.13 | 0.82  (0.78 to 0.85) |

**Table S4 Area under the Receiver Operating Characteristic curve in predicting mortality (in-hospital, 24hour, 48hour, 72hour) at index NEWS2 (or NEWS) ≥ 5 in COVID-19 and Non-COVID-19 emergency medical admissions**

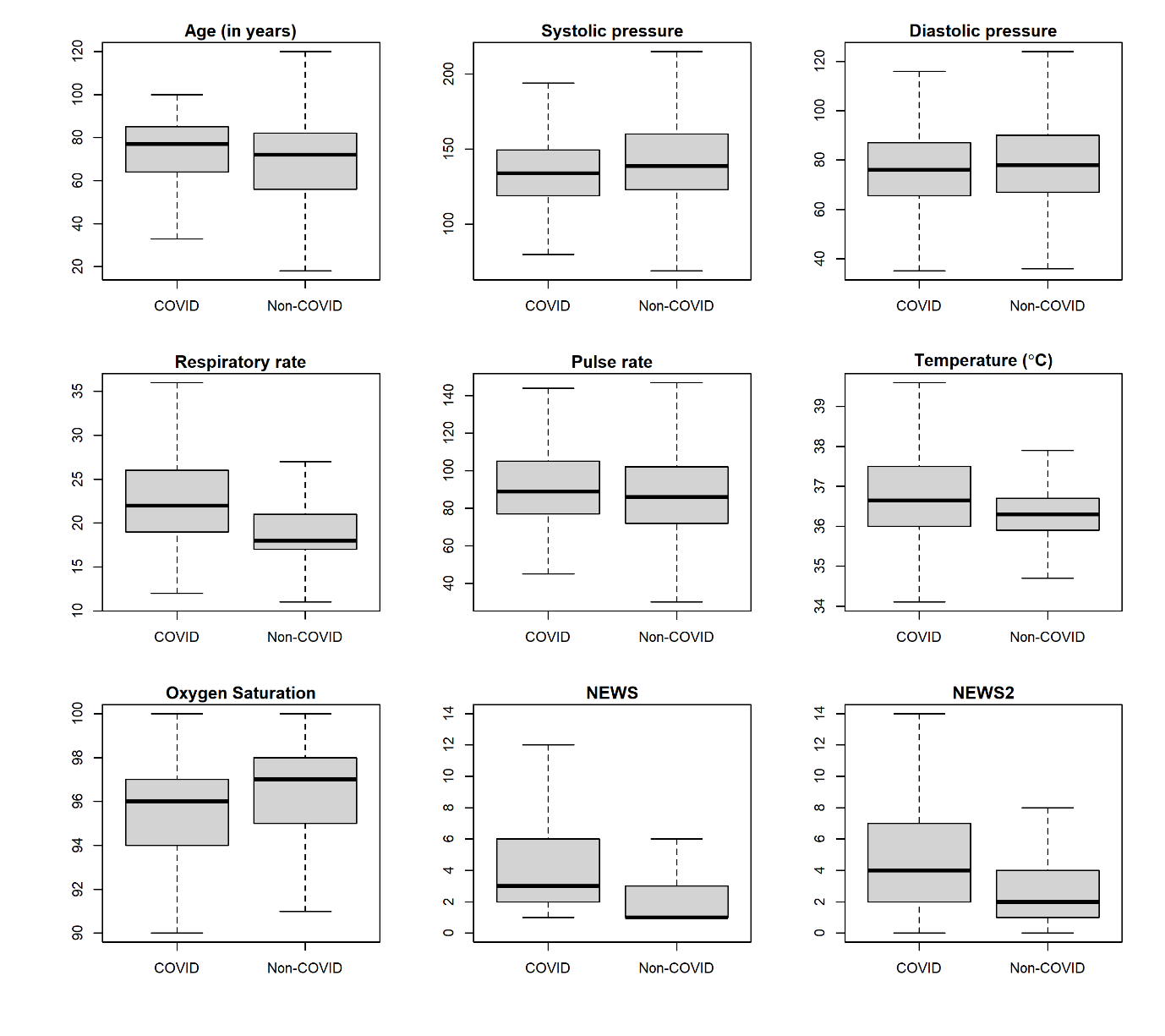

**Figure S1 Boxplots without outliers for continuous covariates for COVID-19 versus Non-COVID-19 admissions**

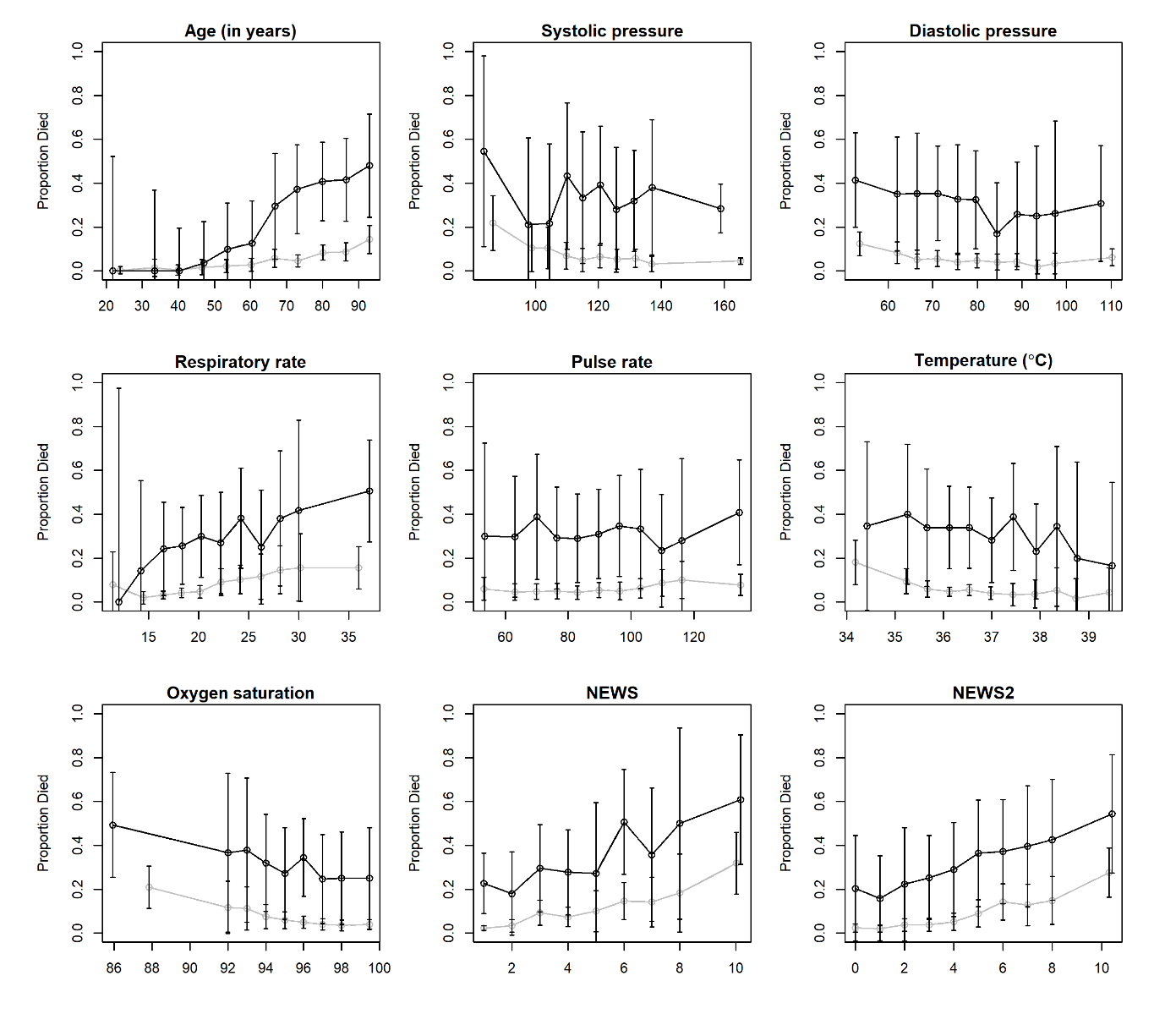

**Figure S2 Line plots showing the observed risk of in-hospital mortality (95% confidence intervals) with continuous covariates for COVID-19 versus Non-COVID-19 admissions**

| Admission type | Medical N= 5313 | | Surgical N=1129 | | Both N=6444 | |
| --- | --- | --- | --- | --- | --- | --- |
|  | COVID+ | COVID- | COVID+ | COVID- | COVID+ | COVID- |
| Number of admission (N) | 588 | 4727 | 32 | 1097 | 620 | 5824 |
| AUC (95% CI) for NEWS | 0.64 (0.60 -0.69) | 0.74 (0.71 -0.77) | 0.63 (0.43 -0.83) | 0.71 (0.63 -0.80) | 0.64 (0.60 -0.69) | 0.74 (0.71 -0.77) |
| AUC (95% CI) for NEWS 2 | 0.64 (0.60 -0.69) | 0.73 (0.70 -0.77) | 0.65 (0.46 -0.85) | 0.74 (0.65 -0.82) | 0.64 (0.60 -0.69) | 0.74 (0.71 -0.77) |

**Table S5: Performance of NEWS and NEWS in predicting the in-hospital mortality in medical and surgical admission.**

*COVID+ = COVID-19 admissions; COVID- = Non-COVID-19 admissions*
